# Monovalent mRNA XBB.1.5 vaccine effectiveness against COVID-19 hospitalization in Quebec, Canada: impact of variant replacement and waning protection during 10-month follow-up

**DOI:** 10.1101/2024.11.13.24317190

**Authors:** Sara Carazo, Danuta M. Skowronski, Nicholas Brousseau, Charles-Antoine Guay, Chantal Sauvageau, Étienne Racine, Denis Talbot, Iulia Gabriela Ionescu, Judith Fafard, Rodica Gilca, Jonathan Phimmasone, Philippe De Wals, Gaston De Serres

## Abstract

**Background:** We evaluated mRNA COVID-19 vaccine effectiveness (VE) for the XBB.1.5 formulation against COVID-19 hospitalizations among adults aged ≥60 years during a ten-month follow-up period.

**Methods:** We conducted a test-negative case-control study using Quebec population-based administrative data. Specimens collected from individuals aged ≥60 years tested at an acute-care hospital from October 2023 to August 2024 were considered test-positive cases if hospitalized for COVID-19, or controls if test-negative for SARS-CoV-2. Vaccination was defined by receipt of at least one mRNA XBB-vaccine (autumn or spring) dose. Multivariate logistic regression analyses estimated VE relative to several comparator groups, primarily those last-vaccinated in 2022, by subvariant predominant period (XBB, JN and KP), by time since XBB-vaccination and by number of XBB-vaccine doses (KP period).

**Results:** Participants overall and by XBB, JN and KP periods included: 5532 (4.9%) test-positive cases (1321, 1838 and 1372, respectively) and 108473 (95.1%) test-negative controls (12881, 53414 and 28595, respectively); 14584 specimens were collected during periods of subvariant cocirculation. By subvariant period, 3322 (25.8%), 27041 (50.6%) and 15401 (53.9%) controls, respectively, were considered XBB-vaccinated. Overall VE was 30% (95%CI:24–35) and by XBB, JN or KP period: 54% (95%CI:46–62), 23% (95%CI:13–32) and 0% (95%CI:-18–15), respectively. During each subvariant period, the hospitalization risk was reduced only during the first four months post-vaccination.

**Conclusions:** Among individuals aged ≥60 years, mRNA XBB-vaccination provided meaningful, albeit limited and short-term, protection against COVID-19 hospitalization due to XBB, JN and KP subvariants in. Better vaccines are needed to effectively reduce COVID-19 disease burden.

**summary:** Based on a population-based test-negative study, XBB.1.5-vaccine effectiveness against COVID-19 hospitalizations was 30% from October 2023 to August 2024 among ≥60-year-olds. By subvariant period, effectiveness was 54% (XBB-period), 23% (JN-period) and 0% (KP-period), with no protection past 5-6 months post-XBB-vaccination.

## INTRODUCTION

SARS-CoV-2 viruses continue to evolve, leading to unpredictable worldwide transitions in lineage predominance, mainly driven by emerging subvariants with features of higher transmissibility and/or immune escape [1]. Vaccine formulations targeting contemporaneous subvariants have been developed to respond to virus evolution. Updated monovalent COVID-19 vaccines targeting the Omicron XBB.1.5 variant (XBB-vaccines) were approved in Canada and the USA in September 2023 [2,3] and administered during 2023 autumn and 2024 spring vaccination campaigns in the province of Quebec, Canada.

Studies evaluating XBB-vaccination have reported early incremental effectiveness (compared to not receiving the XBB-vaccine) of 50-70% to reduce COVID-19 hospitalization risk, but lower protection of 23-54% following replacement of XBB descendants by the Omicron JN.1 sublineage [4–9]. These studies, however, only included the first three to five months following vaccination campaigns and could not inform duration of protection thereafter. In addition, none assessed vaccine effectiveness (VE) against severe outcomes associated with subsequently emergent Omicron KP.2 and KP.3 subvariants, which became the predominant circulating lineages by May 2024 [10].

Using Quebec population-based administrative data, we evaluated effectiveness of mRNA XBB.1.5 vaccination (XBB-vaccination) against hospitalizations for COVID-19 from October 2023 to August 2024 among adults aged ≥60 years overall, by period of subvariant predominance, and by time since XBB-vaccination.

## METHODS

### Study design and population

This population-based test-negative case-control study utilizing specimens tested for SARS-CoV-2 at any acute-care hospital in the province of Quebec, compared the XBB-vaccination status of test-positive cases (COVID-19 hospitalizations) with test-negative controls (hospitalized or not). Tests were included if conducted among adults aged ≥60 years who presented with COVID-19 symptoms from October 29, 2023 (epi-week 2023-44) to August 17, 2024 (epi-week 2024-33). Analyses are specimen-based: individuals could contribute multiple tests to the analysis.

### Procedures

In Quebec, a dose of mRNA XBB-vaccine was recommended for adults aged ≥60 years and other high-risk groups during the autumn campaign commencing September 29, 2023, for long-term care facilities (LTCF) and other congregate living facilities and October 10, 2023 for general population [11]. Influenza and COVID-19 vaccines were offered simultaneously to individuals targeted for both vaccines. A 2024 spring dose of XBB-vaccine (available from April 1, 2024) was recommended for persons aged ≥80 years, those living in LTCF or private homes for older people and immunocompromised individuals [12].

Subvariant identification during the study period was based on whole-genome sequencing of randomly selected specimens among all SARS-CoV-2 positive nucleic acid amplification tests (NAAT). We defined subvariant periods based on >60% predominance reported by these laboratories [13]. In general, XBB subvariants replaced BQ.1 subvariants and were predominant in Quebec starting February 2023. Accordingly, the XBB period spanned from epi-week 2023-44 to 2023-48, during which 69% to 91% of weekly sequenced specimens were XBB and subvariants (18% to 34%) or EG.5 and subvariants (49% to 60%). The JN period spanned from epi-week 2023-52 to 2024-17, during which weekly-sequenced specimens that were identified as BA.2.86, JN.1 or subvariants represented 64% to 100% of all specimens. The KP period spanned from epi-week 2024-20 to 2024-33 (end of the study), during which 61% to 80% of weekly-sequenced specimens were KP.2 and subvariants (5% to 19%), or KP.3 and subvariants (38% to 69%) (Figure 1).

**Figure 1.**
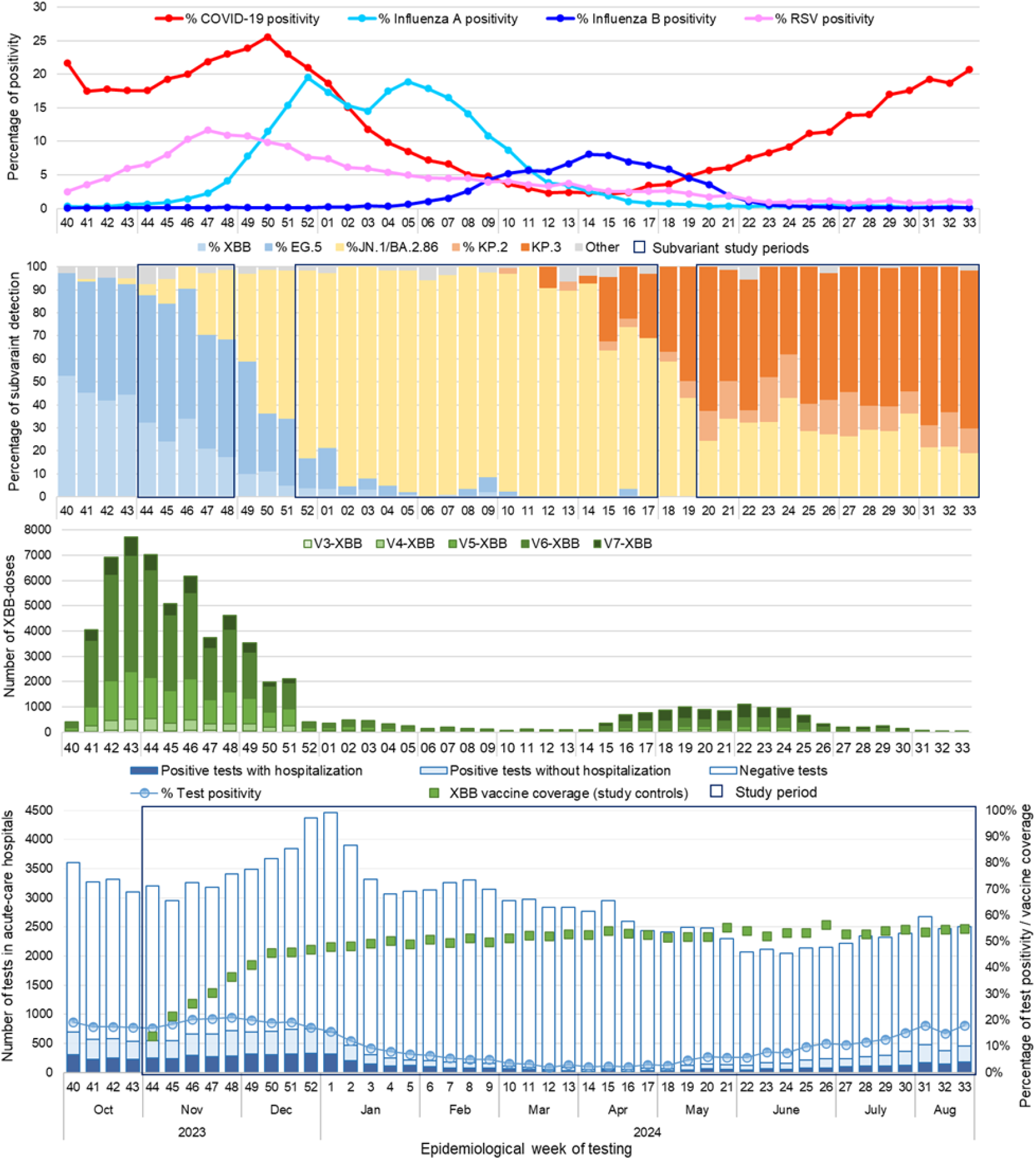
Provincial respiratory viruses and SARS-CoV-2 subvariant circulation, doses of XBB-vaccine administered among study participants, and SARS-CoV-2 tests performed in acute-care hospitals by epidemiological week Abbreviations: V3-XBB to V7-XBB, XBB-vaccine administered as dose 3 to dose 7 Note 1: Subvariants included as XBB/EG.5 detection: XBB, XBB.1.16, XBB.1.5, XBB.1.9, EG.5.1, HK.3, HV.1; as JN.1 detection: JN.1, BA.2.86; as KP.2/3 detection: KP.2, KP.3, KP.3.1.1, KP.3.1.4 Note 2: XBB-vaccine coverage represents at least one dose of XBB-vaccine among test-negative controls

As previously described [14], information on all NAATs for SARS-CoV-2 is centralized in Quebec’s provincial laboratory database. For each NAAT, the reason for testing is documented and, despite some evolution in reporting instructions over time, the code for tests conducted in acute-care hospitals (emergency room or inpatient) due to symptoms compatible with COVID-19 has remained unchanged since 2020. Using a unique personal identifier, we merged this provincial laboratory database with the population-based provincial immunization registry, the hospitalization administrative database and the provincial chronic disease surveillance database [15]. Data were extracted on August 26, 2024.

The primary exposure was defined as receipt of at least one dose of XBB-vaccine administered at any time throughout the study period (autumn and/or spring campaigns). During the KP period, exposure could include one (autumn or spring) or two (autumn and spring) XBB-vaccine doses, assessed in primary analysis combined but also explored separately stratifies by number of XBB-vaccine doses. Three comparison groups were used: a) latest vaccination with an ancestral mRNA monovalent or bivalent vaccine dose received during Quebec’s prior vaccination campaign from July to December 2022 (MV/BV-vaccinated); b) non-XBB vaccination, regardless of the number or timing of previous doses, but excluding individuals having received a non-XBB dose less than 182 days prior to specimen collection (non-XBB-vaccinated); or c) unvaccinated individuals (unvaccinated).

Prior infection was defined as a documented positive SARS-CoV-2 NAAT before October 20, 2023, and ≥90 days before the specimen collection date. We did not have information on the results of rapid antigen detection tests, broadly used since 2022.

COVID-19 hospitalization was defined as a hospital admission during ≥24 hours within 0 to 6 days after a positive SARS-CoV-2 NAAT performed due to COVID-19-compatible symptoms, with COVID-19 being the main reason for admission.

We excluded tests from individuals living in a LTCF because their hospital referral criteria differ from those in the general population, and from immunocompromised individuals identified in the chronic disease surveillance database. We additionally excluded tests: a) of specimens collected within seven days prior to a positive test (probable false negative results), or within 90 days after a positive test (compatible with prolonged SARS-CoV-2 shedding); b) with negative results collected within 15 days after a negative test, to include a single test by respiratory episode; c) of individuals with XBB-vaccination <7 days or non-XBB vaccination <182 days before specimen collection; d) of individuals with invalid vaccination dates or intervals, or who received more than seven vaccine doses or were vaccinated only with non-mRNA vaccines; and e) positive tests done in the emergency room that were not followed by hospital admission.

### Statistical analyses

We used logistic regression models to derive adjusted odds ratios (OR) and their 95% confidence intervals (95% CI) comparing the odds of XBB-vaccination among positive-test hospitalized cases versus negative-test controls. Estimated VE was calculated as (1 – adjusted OR)*100. We estimated incremental XBB-VE when the comparator was the MV/BV-vaccinated group (main analysis) or alternatively the non-XBB-vaccinated group. Absolute XBB-VE was estimated when the comparator was the unvaccinated group.

All models were adjusted for sex, age group (60-69, 70-79 and ≥80 years), residence (home, private homes for older people), epi-week of specimen collection, chronic respiratory disease, chronic heart disease, cancer, obesity, neurological disease and multimorbidity (at least two chronic diseases) [15].

Waning and subvariant-specific estimates were examined by stratifying by subvariant period, time since XBB-vaccination (one- and two-month intervals) and by four-week calendar periods. Other stratified analyses explored the effect of age, the number of XBB doses during the KP period and hybrid prior infection.

Statistical analyses were performed using SAS (version 9.4).

### Ethical considerations

This study was conducted under the legal mandate of the Quebec National Director of Public Health and was approved by the ethics committee of the Centre de Recherche du Centre Hospitalier Universitaire du Québec-Université Laval.

## RESULTS

### Study population

From the 167373 NAAT performed among adults aged ≥60 years at acute-care hospitals during the study period, 53368 (31.9%) tests were excluded: 3575 (2.1%) for individuals living in LTCF and 10863 (6.5%) for immunocompromised, 7621 (4.6%) specimens collected within seven days prior or 90 days after a positive test, 17672 (10.6%) within 15 days after a negative test, and 6058 (3.6%) with an excluded vaccination-test interval, more than seven doses, non-mRNA vaccines or invalid dates (Supplementary_Figure 1). We also excluded 7579 (4.5%) positive-test cases because not hospitalized after ER consultation.

Among the 114005 NAAT included, 5532 (4.9%) were from COVID-19 hospitalized cases: 1321 (23.9%) during the XBB period, 1838 (33.2%) during the JN period, 1372 (24.8%) during the KP period and 1001 (18.1%) during periods of subvariant cocirculation.

Among cases, 2665 (48.2%) were female, the median age was 80 years (interquartile range (IQR): 73-87), 2895 (52.3%) were 80 years or older, and 4230 (76.5%) had multimorbidity. Among controls, 55861 (51.5%) were female, the median age was 77 years (interquartile range (IQR): 69-84), 43790 (40.4%) were 80 years or older, and 77444 (71.4%) presented with multimorbidity. Sex, age and multimorbidity distribution by case and control status were similar in each subvariant period (Table 1).

**Table 1.** Characteristics of test-participants included in each period analysis.

|  | XBB period |  | JN period |  | KP period |  |
| --- | --- | --- | --- | --- | --- | --- |
| Characteristic | Cases | Controls | Cases | Controls | Cases | Controls |
| N | 1321 | 12881 | 1838 | 53414 | 1372 | 28595 |
| **Sex** |  |  |  |  |  |  |
| Female | 645 (48.8) | 6668 (51.8) | 854 (46.5) | 27612 (51.7) | 689 (50.2) | 14563 (50.9) |
| Male | 676 (51.2) | 6213 (48.2) | 984 (53.5) | 25802 (48.3) | 683 (49.8) | 14032 (49.1) |
| **Age, years** |  |  |  |  |  |  |
| Median (IQR) | 81 (74-87) | 77 (69-84) | 79 (73-86) | 77 (69-84) | 80 (74-87) | 77 (69-84) |
| 60-69 | 185 (14.0) | 3438 (26.7) | 302 (16.4) | 14091 (26.4) | 192 (14.0) | 7344 (25.7) |
| 70-79 | 415 (31.4) | 4223 (32.8) | 623 (33.9) | 17626 (33.0) | 444 (32.4) | 9860 (34.5) |
| ≥80 | 721 (54.6) | 5220 (40.5) | 913 (49.7) | 21697 (40.6) | 736 (53.6) | 11391 (39.8) |
| **Place of residence** |  |  |  |  |  |  |
| Home | 966 (73.1) | 10150 (78.8) | 1488 (81.0) | 42591 (79.7) | 1118 (81.5) | 23457 (82.0) |
| Private homes for older people | 336 (25.4) | 2462 (19.1) | 316 (17.2) | 9678 (18.1) | 236 (17.2) | 4543 (15.9) |
| Other | 19 (1.4) | 269 (2.1) | 34 (1.8) | 1145 (2.1) | 18 (1.3) | 595 (2.1) |
| **Chronic conditions** |  |  |  |  |  |  |
| Multimorbidity <sup>a</sup> | 1029 (77.9) | 9360 (72.7) | 1377 (74.9) | 38166 (71.5) | 1055 (76.9) | 20180 (70.6) |
| Chronic heart disease | 694 (52.5) | 5970 (46.3) | 879 (47.8) | 23847 (44.6) | 663 (48.3) | 12517 (43.8) |
| Chronic lung disease | 474 (35.9) | 4722 (36.7) | 649 (35.3) | 19837 (37.1) | 479 (34.9) | 10442 (36.5) |
| Cancer | 275 (20.8) | 2606 (20.2) | 374 (20.3) | 10563 (19.8) | 283 (20.6) | 5855 (20.5) |
| Neurologic disease / dementia | 245 (18.5) | 1823 (14.2) | 299 (16.3) | 6984 (13.1) | 206 (15.0) | 3444 (12.0) |
| Obesity | 146 (11.1) | 1549 (12.0) | 197 (10.7) | 6015 (11.3) | 162 (1.8) | 3143 (11.0) |
| **Documented infection prior to vaccination campaign** | 218 (16.5) | 2760 (21.4) | 317 (17.2) | 12042 (22.5) | 250 (18.2) | 6740 (23.6) |
| **Number of booster doses received (total vaccine doses)** |  |  |  |  |  |  |
| None (V0-V2) | 142 (10.7) | 1463 (11.4) | 212 (11.5) | 5738 (10.7) | 120 (8.7) | 2996 (10.5) |
| One (V3) | 231 (17.5) | 2336 (18.1) | 310 (16.9) | 8467 (15.9) | 204 (14.9) | 4430 (15.5) |
| Two (V4) | 270 (20.4) | 2769 (21.5) | 313 (17.0) | 8511 (15.9) | 195 (14.2) | 4360 (15.2) |
| Three (V5) | 439 (33.2) | 3588 (27.9) | 388 (21.1) | 11756 (22.0) | 291 (21.2) | 5806 (20.3) |
| Four (V6) | 206 (15.6) | 2369 (18.4) | 501 (27.3) | 15858 (29.7) | 417 (30.4) | 8527 (29.8) |
| Five (V7) | 33 (2.5) | 356 (2.8) | 114 (6.2) | 3084 (5.8) | 145 (10.6) | 2476 (8.7) |
| **Type of last booster dose (total vaccine doses)** |  |  |  |  |  |  |
| Ancestral monovalent (V3-V6) | 482 (36.5) | 4425 (34.4) | 494 (26.9) | 12959 (24.3) | 313 (22.8) | 6543 (22.9) |
| Bivalent (BA.1 or BA.4/5) (V3-V7) | 496 (37.5) | 3671 (28.5) | 318 (17.3) | 7676 (14.4) | 158 (11.5) | 3655 (12.8) |
| Monovalent XBB.1.5 (V3-V7) | 201 (15.2) | 3322 (25.8) | 814 (44.3) | 27041 (50.6) | 781 (56.9) | 15401 (53.9) |
| **Interval in months between last dose and specimen collection, median (IQR)** |  |  |  |  |  |  |
| Any vaccine type | 13.4 (10-19) | 13.1 (1-19) | 12.2 (3-21) | 5.6 (3-21) | 9.1 (8-23) | 9.0 (7-25) |
| Ancestral monovalent / Bivalent | 13.2 (12-14) | 13.2 (12-14) | 15.4 (14-17) | 16.3 (15-18) | 20.9 (20-22) | 20.6 (19-22) |
| Monovalent XBB 1.5 | 0.6 (0-1) | 0.7 (0-1) | 2.5 (2-4) | 3.3 (2-5) | 8.0 (7-9) | 7.2 (6-8) |
| **2023/24 seasonal influenza vaccination (≥14 days before specimen collection)** | 165 (12.5) | 2549 (19.8) | 796 (43.3) | 25414 (47.6) | 731 (53.3) | 13997 (48.9) |

Abbreviations: IQR, interquartile range; V0 to V7, vaccine dose 0 to 7
<sup>a</sup> At least two chronic conditions among the following: chronic respiratory disease, hypertension, cardiovascular disease, neurological disorder, anaemia, diabetes, hypothyroidism, fluid and electrolyte disorders, cancer, kidney disease, obesity, psychosis, liver disease, coagulopathy, weight loss, drug abuse, alcohol abuse, ulcer, paralysis.
Note: Data are n and column percentage except otherwise indicated

By successive XBB, JN and KP subvariant period, 3322 (25.8%), 27041 (50.6%) and 15401 (53.9%) controls were XBB-vaccinated, respectively. Vaccination was associated with older age and multimorbidity (Table 1, Supplementary_Tables 1-3). Among controls, median time since XBB-vaccination increased by successive subvariant period, from 22 days (IQR: 15-32), to 3 months (IQR: 2-5), to 7 months (IQR: 6-8), respectively. Control XBB-vaccine coverage during the last week of March 2024, before the 2024 spring XBB-vaccination campaign, was 52.8% overall: 37.5%, 53.5% and 61.9%, respectively, for 60-69, 70-79 and ≥80 years. The latter estimates are comparable to population vaccine coverage on April 1, 2024 among Quebec adults: 36.4%, 54.2% and 60.7%, respectively, for 60-69, 70-79 and ≥80 years (Figure 1) [16]. Control vaccine coverage of the spring dose at the end of the study period (August 2024, week 33) was 10.4%, among whom 70.7% had also received an autumn XBB-vaccine dose. A similar proportion of XBB-vaccinated cases (1636 (77.0%)) and controls (40463 (77.6%)) had also received an influenza vaccine (Supplementary_Tables 1-3).

In each subvariant period, a higher proportion of controls than cases had a documented NAAT-confirmed SARS-CoV-2 infection prior to the study period (Table 1). A recent documented infection that occurred between May and October 2023 was more frequent among MV/BV-vaccinated (16%) than among XBB-vaccinated participants (7%), suggesting a possible association between recent infection and delayed vaccination (Supplementary_Figure 2).

### Estimated VE

Relative to MV/BV-vaccination, global XBB-VE against COVID-19 hospitalization was 30% (95%CI:24–35): highest during the XBB period at 54% (95%CI:46–62), lower during the JN period at 23% (95%CI:13–32), and with no protection observed during the KP period (VE=0%; 95%CI:-17–15), when 9.7% of XBB-vaccinated had received 2 doses (Table 2). Estimated VE was similar when compared with non-XBB-vaccinated or with unvaccinated participants, suggesting lack or low residual protection from vaccination prior to the 2023 autumn campaign.

**Table 2.** XBB-vaccine effectiveness against COVID-19 hospitalization relative to different comparison groups, by subvariant period.

| Variant period and comparison groups | Cases | Controls | Unadjusted VE<br>(95% CI) | Adjusted VE<br>(95% CI) |
| --- | --- | --- | --- | --- |
| **Global (W2023-44 to W2024-33)** |  |  |  |  |
| XBB-vaccinated | 2126 | 52135 |  |  |
| vs MV\BV-vaccinated in 2022 | 1356 | 20343 | 38.8 (34.4 to 43.0) | 29.9 (24.4 to 35.0) |
| vs non-XBB vaccinated | 3406 | 56338 | 32.5 (28.7 to 36.2) | 29.6 (25.3 to 33.6) |
| vs unvaccinated | 263 | 4273 | 33.7 (24.4 to 41.9) | 35.8 (26.3 to 44.1) |
| **XBB period (W2023-44 to W2023-48)** |  |  |  |  |
| XBB-vaccinated | 201 | 3322 |  |  |
| vs MV\BV-vaccinated in 2022 | 556 | 4384 | 52.3 (43.6 to 59.6) | 54.4 (45.9 to 61.6) |
| vs non-XBB vaccinated | 1120 | 9559 | 48.4 (39.7 to 55.8) | 54.0 (46.1 to 60.7) |
| vs unvaccinated | 62 | 562 | 45.2 (26.1 to 59.3) | 53.8 (37.3 to 66.0) |
| **JN period (W2023-52 to W2024-17)** |  |  |  |  |
| XBB-vaccinated | 814 | 27041 |  |  |
| vs MV\BV-vaccinated in 2022 | 363 | 9069 | 24.8 (14.7 to 33.7) | 22.7 (12.1 to 32.0) |
| vs non-XBB vaccinated | 1024 | 26373 | 22.5 (14.9 to 29.4) | 25.2 (17.5 to 32.1) |
| vs unvaccinated | 87 | 2055 | 28.9 (10.9 to 43.3) | 30.8 (12.7 to 45.1) |
| **KP period (W2024-20 to W2024-33)** |  |  |  |  |
| XBB-vaccinated | 781 | 15401 |  |  |
| vs MV\BV-vaccinated in 2022 | 204 | 4299 | -6.9 (-25.1 to 8.7) | -0.5 (-16.7 to 15.2) |
| vs non-XBB vaccinated | 591 | 13194 | -13.2 (-26.3 to -1.5) | -0.7 (-12.8 to 10.1) |
| vs unvaccinated | 52 | 1088 | -6.1 (-41.4 to 20.4) | 7.4 (-24.2 to 30.9) |

Abbreviations: CI, confidence interval; MV/BV, ancestral monovalent or bivalent booster dose received between July and December 2022; VE, vaccine effectiveness; W, week
Note: Logistic regression model adjusted for sex, age group, chronic conditions, place of residence and epi-week

Stratification for time since XBB-vaccination showed that significant protection against hospitalization was only observed during the first four months post-vaccination for each subvariant period (Figure 2, Supplementary_Table 4). It was higher during the first two months post-vaccination during XBB (VE=55%; 95%CI:46–62), JN (VE=23%; 95%CI:9-36) and KP periods (VE=60%; 95%CI:39–74). As XBB predominance ended epi-week 2023-48, time-since-vaccination beyond two months could not be evaluated. During the JN period, observed VE was 17% (95%CI:-9-36) at five to six months and -5% (95%CI:-160-57) at seven months post-vaccination. During the KP period, effectiveness decreased with time reaching 3% at seven to eight months post-vaccination. After standardisation for time since vaccination during the KP period, a second XBB-dose was associated with similar protection to a single XBB-vaccination through the 4^th^ month post-vaccination, mainly reflecting spring doses (Table 3).

**Table 3.**
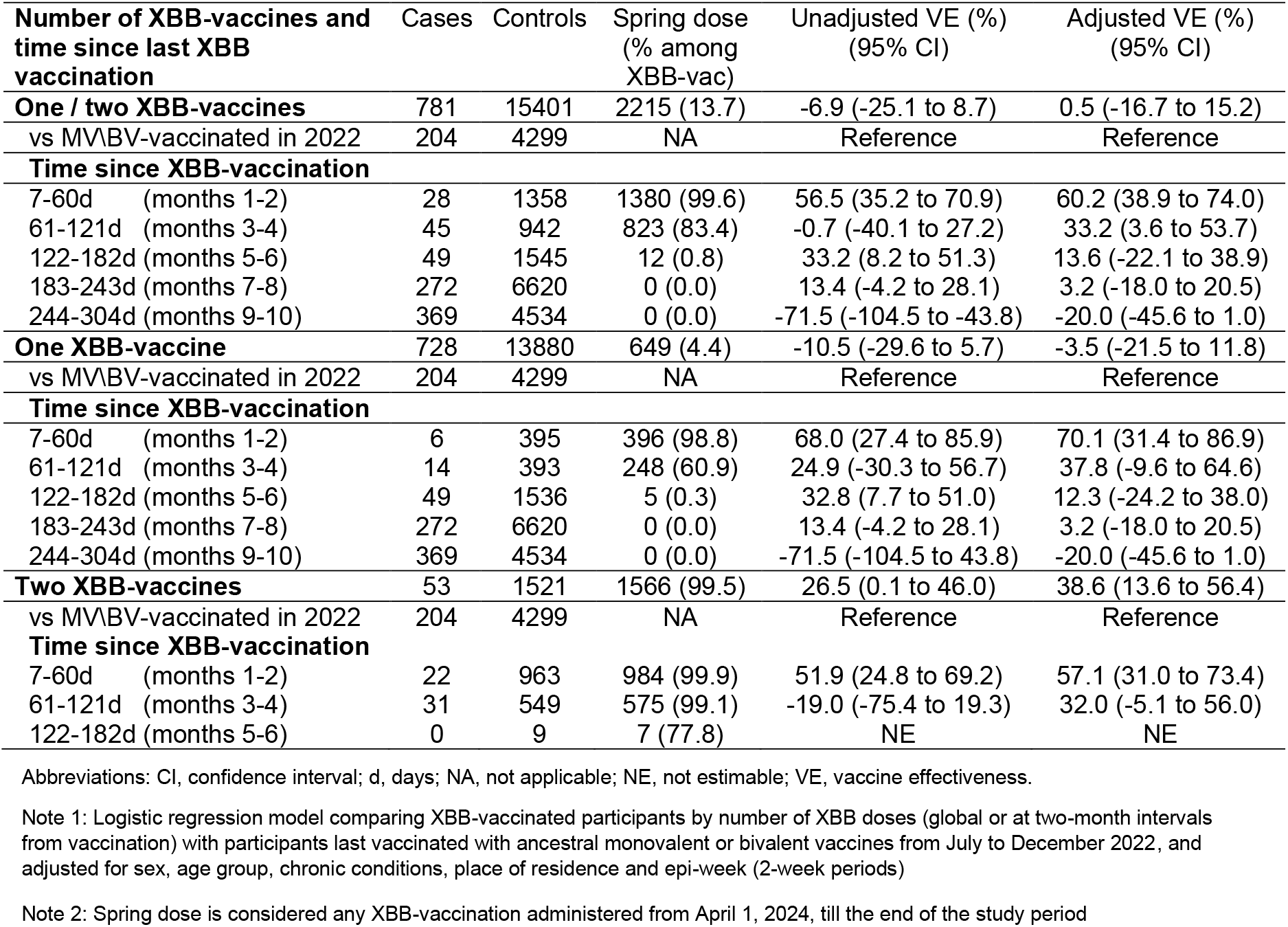
XBB vaccine effectiveness against COVID-19 hospitalization relative to monovalent or bivalent vaccination in 2022, by number of XBB-doses during the KP period.

When stratifying by calendar time, XBB-vaccination was associated with a 50% reduction in COVID-19 hospitalization risk during the first eight weeks of the study period (November and December 2023), decreasing to 30% in January 2024 and to no protection from February to August 2024 (Figure 3). This rapid loss of protection was contemporaneous with increased median time since vaccination, subvariant replacement, and low uptake of the spring dose (Figure 1).

**Figure 2:**
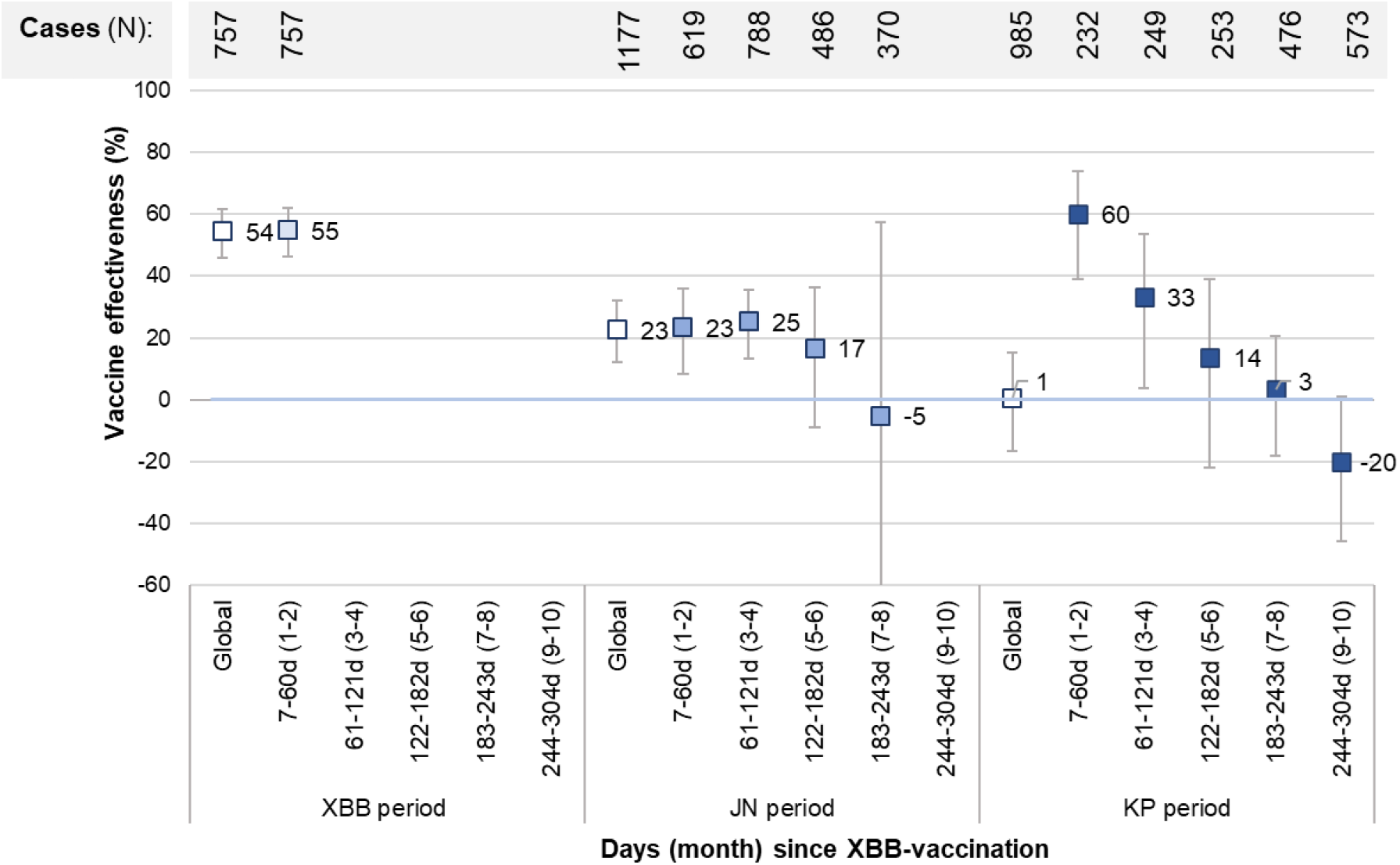
XBB-vaccine effectiveness against COVID-19 hospitalization relative to monovalent or bivalent vaccination in 2022, by time since XBB-vaccination and variant period Note: Logistic regression model comparing XBB-vaccinated participants at two-month intervals from vaccination with participants last vaccinated with an ancestral monovalent or a bivalent vaccine from July to December 2022 and adjusted for sex, age group, chronic conditions, place of residence and epi-week (2-week periods for the time since vaccination analyses)

**Figure 3:**
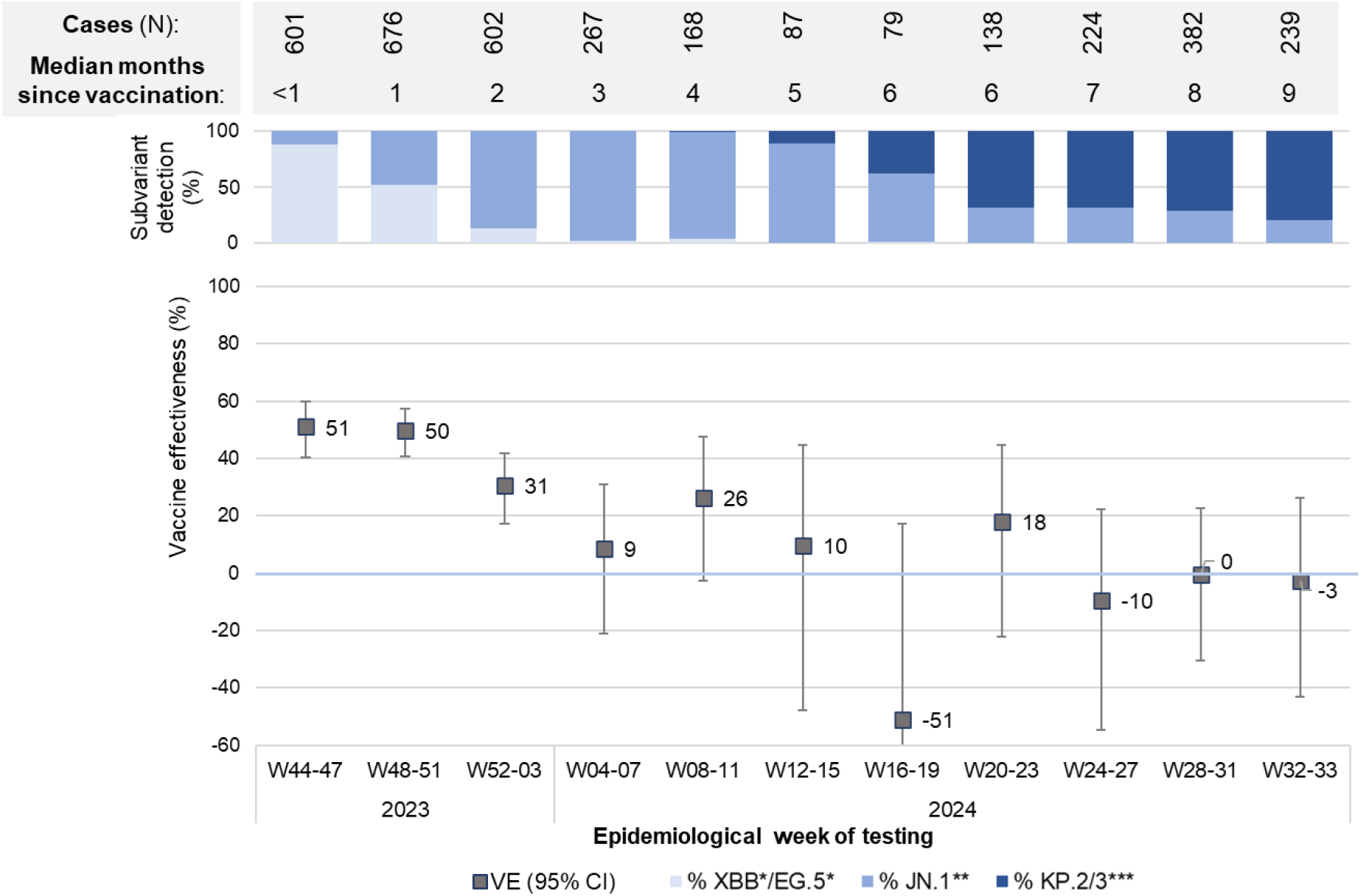
XBB-vaccine effectiveness against COVID-19 hospitalization relative to monovalent or bivalent vaccination in 2022, by calendar time (four epi-week periods) ^*^ XBB, XBB.1.5, XBB.1.9, XBB.1.16, eG.5.1, HK.3, HV.1 ^**^ JN.1, BA.2.86 ^***^ KP.2, KP.3, KP.3.1.1, KP.3.1.4 Note: Logistic regression model comparing XBB-vaccinated participants with participants last vaccinated with an ancestral monovalent or a bivalent vaccine from July to December 2022 and adjusted for sex, age group, chronic conditions, place of residence and two epi-week periods

In subgroup analyses, XBB-vaccination among individuals with prior NAAT-confirmed infection compared to MV/BV-vaccinated regardless prior infection was associated with higher point estimates of VE than main global estimates during each subvariant period (74%, 41% and 6% vs. 54%, 23% and 0%, respectively) (Supplementary_Table 5). Overall and subvariant-specific VE estimates were unexpectedly lower for 60-to 69-year-olds than for older groups, but differences decreased in analyses restricted to the first two months post-XBB-vaccination (Supplementary_Table 6).

## DISCUSSION

During a ten-month period of follow-up, XBB-vaccinated adults aged 60 years or older had an estimated 30% reduction in their risk of COVID-19 hospitalization. Restricted to the first two months post-vaccination, VE was 55% during the XBB period, 23% during the JN period and 60% during the KP period, but effectiveness waned over time, with minimal to no protection demonstrated beyond fourth months, regardless of the subvariant.

Prior published studies have reported relative XBB-VE of 53% to 67% against hospitalization related to XBB lineages. These estimates mostly reflect short-term protection, as XBB/EG.5 subvariant predominance was overtaken by JN subvariants by the end of 2023 [5,6,8,10,17,18]. Studies evaluating VE against hospitalizations during both XBB and JN periods showed lower relative protection of 35% to 54% against JN sublineages [4,5,9,19], persisting when comparing similar intervals since vaccination. Waning protection during the JN period was not observed in most studies reporting a four- or five-month post-vaccination follow-up [7,9,20,21], while one study reported waning protection against JN-lineage hospitalization from 33% at <3 months post-vaccination to 23% at 3-5 months [5].

Our data are consistent with the literature for XBB and early JN periods, but our ten-month follow-up spanning the spring 2024 dose allowed us to show that shortly after vaccination, VE was similarly high against XBB (55%) and KP (60%) sublineages, with lower estimate against JN (23%) sublineage. It declined over the following few months with no protection found past five to six months post-XBB-vaccination. This suggests that waning of vaccine immunity and not sublineage shift accounts for most of the observed loss of protection over time. The similarity of XBB-VE estimates with each of the three comparator groups (MV/BV-vaccinated, non-XBB-vaccinated and unvaccinated) also suggests that there was absence or little residual protection in individuals vaccinated >6 months prior to the autumn 2023 campaign, as reported in studies from the USA and England [8,19].

Our study has several limitations. The lack of individual-level genome sequencing might have led to some misclassification in the sublineage-specific estimates, mainly during the KP period where 20% to 30% of analyzed specimens were JN sublineages. Residual confounding is possible if unmeasured determinants of vaccine-seeking behaviour are associated with COVID-19 testing and hospitalization risk. Among our participants with documented prior infection (∼20%), a SARS-CoV-2 infection in September or October 2023 was twice as frequent among MV/BV-vaccinated than among XBB-vaccinated. VE would be underestimated if individuals with increased immunity due to a recent SARS-CoV-2 infection had delayed or avoided XBB-vaccination [22]. Its impact should be small in the context of limited proportion with recent infections and myriad other reasons for non-vaccination but may partially explain the unexpected high VE within two months post-vaccination during the KP period, where delayed (spring) XBB-vaccine receipt as first XBB-dose might have been associated with a recent prior infection. Hybrid immunity conferred by prior infection and vaccination have been previously associated with the highest protection against COVID-19 severe outcomes [14,23]. In the context where most mild COVID-19 infections are no longer documented or NAAT-confirmed, and most people have experienced at least one prior infection, our overall results reflect the incremental XBB-VE over the current (but unknown) population immunity conferred by prior infection. We adjusted for the main determinants of COVID-19 hospitalizations, but we did not have information on factors such as ritonavir-boosted nirmatrevil use, frailty, race and ethnicity which might influence vaccination uptake and disease severity. Depletion-of-susceptible bias may arise when infection rates differ between vaccinated and unvaccinated populations, potentially creating a spurious decrease in VE over time [24]. Although vaccination status might influence contacts especially within certain high-risk groups, the impact of this bias alone could not explain the observed waning, as shown in previous epidemiological studies [25]. XBB and influenza vaccine coadministration might potentially underestimate XBB-VE in test-negative designs [26,27]. Even if we did not have information on influenza results to evaluate its impact, the circulation of SARS-CoV-2 and influenza overlapped only during the JN period in Quebec [28], and we found a similar proportion of influenza-vaccinated among XBB-vaccinated cases and controls, suggesting a limited impact of this bias on our conclusions. Despite these limitations, our conclusions seem robust as shown by results using different comparators and by the time-dependant waning gradient for both JN and KP periods acknowledging wide CI.

This end-of-season study presents a comprehensive portrait of the evolution of the protection conferred by XBB-vaccination at a population level, first reporting sublineage-specific estimates during predominant KP circulation and the first VE estimates associated with two XBB-vaccine doses. Our results align with other reported XBB-VE estimates up to five-months post-vaccination [4,5,7,19] and extends them to a ten-month follow-up including three predominant circulating sublineages finding minimal to no protection beyond the fourth post-vaccination month. Further studies will be necessary to corroborate our conclusions. Moreover, our study leads to several important public health considerations related to ongoing COVID-19 vaccination programs including their timing; their likelihood of success in the context of suboptimal VE, short duration of protection, and ultimately vaccination fatigue; and the need to pursue better COVID-19 vaccines. Unlike influenza, COVID-19 is not a seasonal winter disease. Worldwide, each new COVID-19 wave is linked to a variant shift regardless of the time of the year [1,10]. Therefore, timing COVID-19 vaccination campaigns along that for influenza might be operationally preferable, but not yet optimal with respect to impact. With only short-lasting protection, vaccination only once per year leaves individuals unprotected for much of the time, but more doses lead to vaccine fatigue, with current vaccine coverage as low as 14% for the spring dose among ≥80-year-olds [16]. Finally, despite their high VE and remarkable impact at the beginning of the pandemic, we are now at a stage where mRNA vaccines are showing only modest protection lasting a few months against even severe outcomes like hospitalization, with a recurring need for update to address the emergence of new variants. While exploring the reasons for this dramatic change in the performance of mRNA vaccines we urgently need to determine if other vaccine technologies may yield better and longer-lasting benefits in the current context.

In the meantime, we conclude from our observations that the XBB formulation of mRNA COVID-19 vaccines provided meaningful, albeit limited and short-lived, protection against hospitalization due to XBB, JN and KP sublineages in individuals aged ≥60 years.

## ACKNOWLEDGMENTS

We would like to thank Ella Diendere (Institut national de santé publique du Québec) for her support to provide data on SARS-CoV-2 lineages circulation in Québec.

## Financial support

This work was supported by the Ministère de la santé et des services sociaux du Québec.

### Author contributions

All authors had full access to all of the data in the study and take responsibility for the integrity of the data and the accuracy of the data analysis. **Concept and design**: SC, DMS, GDS. **Acquisition, analysis, or interpretation of data**: All authors. **Drafting of the manuscript**: SC. **Critical revision of the manuscript for important intellectual content**: All authors. **Statistical analysis**: SC, JP. **Supervision**: GDS

### Data availability

Individual data not publicly available

