## Supplemental material for "Monovalent mRNA XBB.1.5 vaccine effectiveness against COVID-19 hospitalization in Quebec, Canada: impact of variant replacement and waning protection during 10-month follow-up"

**Supplementary Material**

**Supplementary Figure 1:** Flowchart of study population……………………………….3

**Supplementary Figure 2:** Temporal distribution of first and second positive SARS-CoV-2 nucleic acid amplification test prior to XBB-vaccination campaign, by participant vaccination status……………………………………………………………………….…4

**Supplementary Table 1**. Characteristics of study participants during XBB period by outcome and vaccination status……………………………………………………….....5

**Supplementary Table 2**. Characteristics of study participants during JN period by outcome and vaccination status………………………………………………………..7

**Supplementary Table 3**. Characteristics of study participants during KP period by outcome and vaccination status…………………………………………………..……9

**Supplementary Table 4.** XBB-vaccine effectiveness against COVID-19 hospitalization by time since XBB-vaccination and subvariant period……….……………………………11

**Supplementary Table 5.** XBB-vaccine effectiveness against COVID-19 hospitalization restricted to participants with prior NAAT-confirmed infection, by subvariant period..….12

**Supplementary Table 6.** XBB-vaccine effectiveness against COVID-19 hospitalization, by age group and subvariant period……………………………………………………….13

**Supplementary Figure 1:** Flowchart of study population

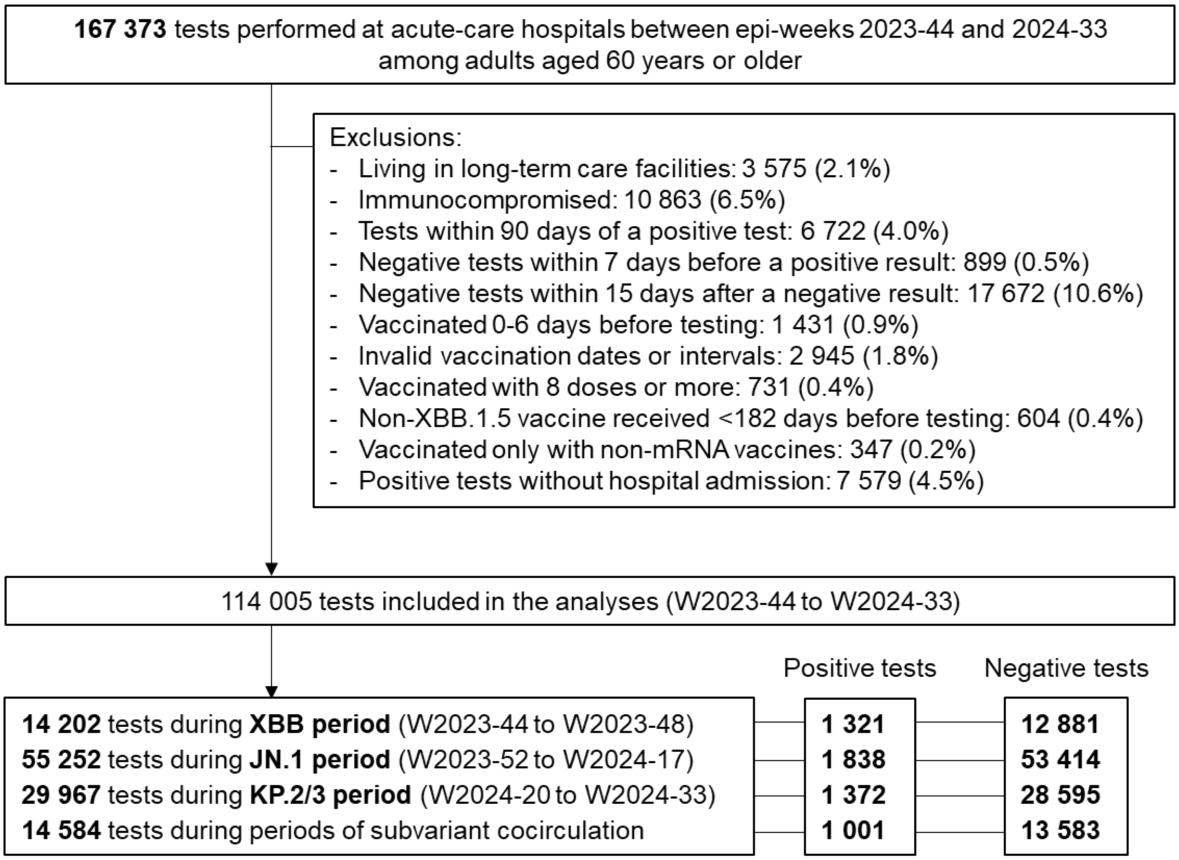

**Supplementary Figure 2:** Temporal distribution of first and second positive SARS-CoV-2 nucleic acid amplification test prior to XBB-vaccination campaign, by participant vaccination status

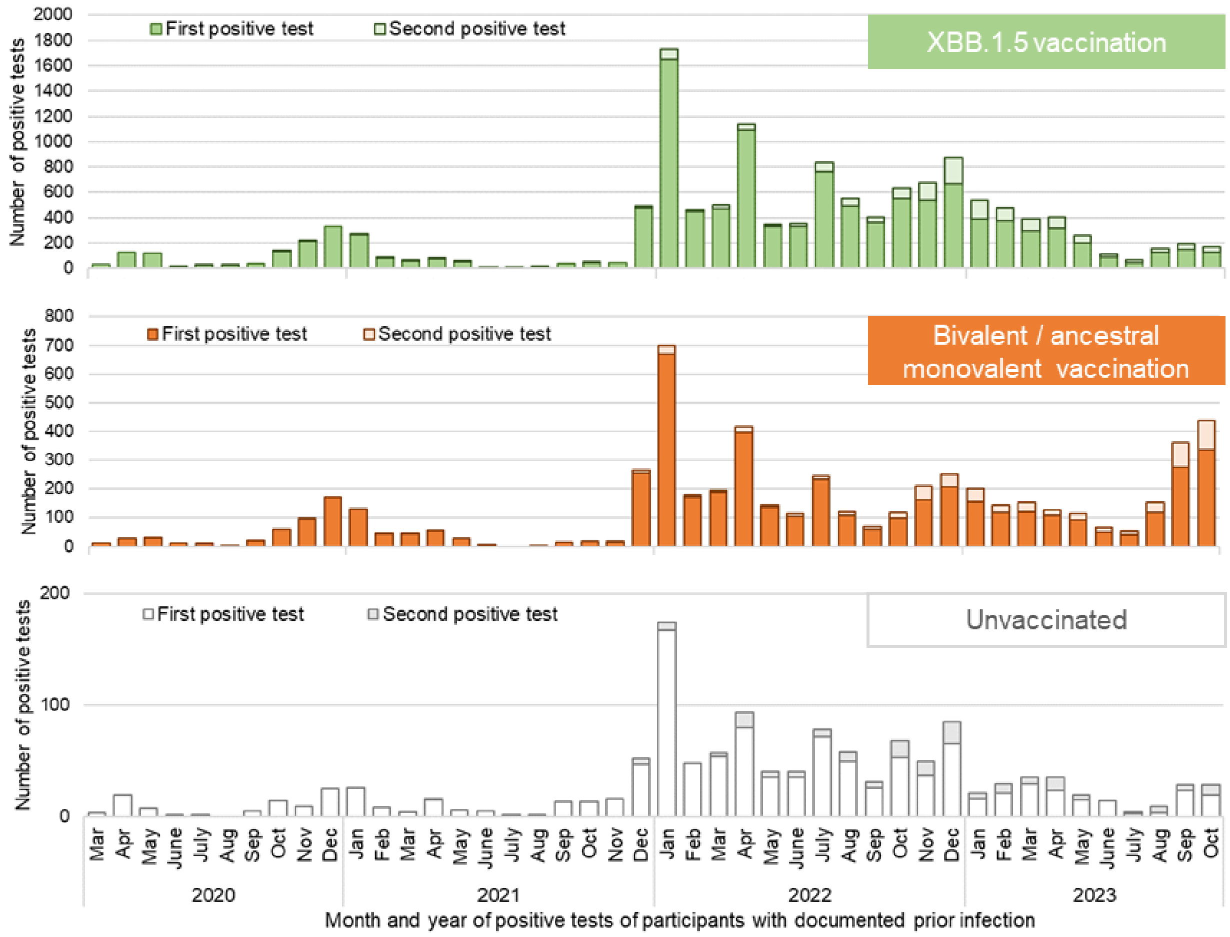

**Supplementary Table 1**. Characteristics of study participants during XBB period by outcome and vaccination status

| XBB period | Cases | | Controls | |
|---|---|---|---|---|
| **Characteristic** | **XBB vaccinated** | **MV/BV vaccinated** | **XBB vaccinated** | **MV/BV vaccinated** |
| **N** | 201 | 556 | 3322 | 4384 |
| **Sex** | | | | |
| Female | 89 (44.3) | 277 (49.8) | 1755 (52.8) | 2285 (52.1) |
| Male | 112 (55.7) | 279 (50.2) | 1567 (47.2) | 2099 (47.9) |
| **Age, years** | | | | |
| Mean (standard deviation) | 82 (9.3) | 81 (8.6) | 79 (9.3) | 78 (9.6) |
| 60-69 | 21 (10.4) | 55 (9.9) | 664 (20.0) | 1011 (23.1) |
| 70-79 | 63 (31.3) | 180 (32.4) | 1129 (34.0) | 1443 (32.9) |
| ≥80 | 117 (58.2) | 321 (57.7) | 1529 (46.0) | 1930 (44.0) |
| **Place of residence** | | | | |
| Home | 152 (75.6) | 380 (68.3) | 2455 (73.9) | 3284 (74.9) |
| Private homes for older people | 49 (24.4) | 165 (29.7) | 805 (24.2) | 984 (22.4) |
| Other | 0 (0.0) | 11 (2.0) | 62 (1.9) | 116 (2.6) |
| **Chronic conditions** | | | | |
| At least two chronic conditions [a] | 174 (86.6) | 432 (77.7) | 2530 (76.2) | 3235 (73.8) |
| Chronic heart disease | 126 (62.7) | 300 (54.0) | 1608 (48.4) | 2082 (47.5) |
| Chronic lung disease | 80 (39.8) | 188 (33.8) | 1313 (39.5) | 1575 (35.9) |
| Cancer | 49 (24.4) | 121 (21.8) | 767 (23.1) | 931 (21.2) |
| Neurologic disease / dementia | 40 (19.9) | 114 (20.5) | 505 (15.2) | 627 (14.3) |
| Obesity | 22 (10.9) | 54 (9.7) | 412 (12.4) | 519 (11.8) |
| **Documented prior infection history** | 28 (13.9) | 103 (18.5) | 737 (22.2) | 929 (21.2) |
| **Interval in months between prior infection and specimen collection,** median (IQR) | 16.1 (13-20) | 18.7 (12-22) | 16.0 (11-21) | 16.2 (10-22) |
| **Number of booster doses received** | | | | |
| One (V3) | 1 (0.5) | 16 (2.9) | 39 (1.2) | 122 (2.8) |
| Two (V4) | 10 (5.0) | 172 (30.9) | 199 (6.0) | 1722 (39.3) |
| Three (V5) | 44 (21.9) | 365 (65.6) | 767 (23.1) | 2519 (57.5) |
| Four (V6) | 113 (56.2) | 3 (0.5) | 1966 (59.2) | 21 (0.5) |
| Five (V7) | 33 (16.4) | 0 (0.0) | 351 (10.6) | 0 (0.0) |

| | | | | |
|---|---|---|---|---|
| **Interval in months between last dose and specimen collection,** median (IQR) | 0.6 (0.4-1) | 13.2 (12-14) | 0.7 (0.5-1) | 13.2 (12-14) |
| **2023/24 seasonal influenza vaccination** (≥14 days before specimen collection) | 107 (53.2) | 25 (4.5) | 2120 (63.8) | 245 (5.6) |

Abbreviations: V3 to V7, total number of vaccine doses 3 to 7; MV/BV, monovalent or bivalent booster dose received between July and December 2022; IQR, interquartile range

[a] At least two chronic conditions among the following: chronic respiratory disease, hypertension, cardiovascular disease, neurological disorder, anaemia, diabetes, hypothyroidism, fluid and electrolyte disorders, cancer, kidney disease, obesity, psychosis, liver disease, coagulopathy, weight loss, drug abuse, alcohol abuse, ulcer, paralysis.

Note: Numbers are n and column proportions except otherwise indicated

**Supplementary Table 2**. Characteristics of study participants during JN period by outcome and vaccination status

| JN period | Cases | | Controls | |
|---|---|---|---|---|
| **Characteristic** | **XBB vaccinated** | **MV/BV vaccinated** | **XBB vaccinated** | **MV/BV vaccinated** |
| **N** | 814 | 363 | 27041 | 9069 |
| **Sex** | | | | |
| Female | 387 (47.5) | 176 (48.5) | 14141 (52.3) | 4568 (50.4) |
| Male | 427 (52.5) | 187 (51.5) | 12900 (47.7) | 4501 (49.6) |
| **Age, years** | | | | |
| Mean (standard deviation) | 80 (9.0) | 79 (8.9) | 79 (9.3) | 77 (9.6) |
| 60-69 | 111 (13.6) | 59 (16.3) | 4986 (18.4) | 2520 (27.8) |
| 70-79 | 271 (33.3) | 129 (35.5) | 9057 (33.5) | 3040 (33.5) |
| ≥80 | 432 (53.1) | 175 (48.2) | 12998 (48.1) | 3509 (38.7) |
| **Place of residence** | | | | |
| Home | 597 (73.3) | 296 (81.5) | 19523 (72.2) | 7348 (81.0) |
| Private homes for older people | 198 (24.3) | 62 (17.1) | 6836 (25.3) | 1525 (16.8) |
| Other | 19 (2.3) | 5 (1.4) | 682 (2.5) | 196 (2.2) |
| **Chronic conditions** | | | | |
| At least two chronic conditions [a] | 631 (77.5) | 272 (74.9) | 20466 (75.7) | 6418 (70.8) |
| Chronic heart disease | 408 (50.1) | 161 (44.4) | 13045 (48.2) | 3910 (43.1) |
| Chronic lung disease | 309 (38.0) | 128 (35.3) | 10538 (39.0) | 3284 (36.2) |
| Cancer | 190 (23.3) | 69 (19.0) | 5895 (21.8) | 1802 (19.9) |
| Neurologic disease / dementia | 164 (20.1) | 62 (17.1) | 4014 (14.8) | 1108 (12.2) |
| Obesity | 85 (10.4) | 45 (12.4) | 3157 (11.7) | 1034 (11.4) |
| **Documented prior infection history** | 145 (17.8) | 58 (16.0) | 5946 (22.0) | 2262 (24.9) |
| **Interval in months between prior infection and specimen collection,** median (interquartile range) | 18.5 (14-23) | 17.6 (6-24) | 18.9 (14-24) | 13.8 (5-24) |
| **Number of booster doses received** | | | | |
| One (V3) | 14 (1.7) | 16 (4.4) | 309 (1.1) | 309 (3.4) |
| Two (V4) | 48 (5.9) | 151 (41.6) | 1822 (6.7) | 4362 (48.1) |
| Three (V5) | 168 (20.6) | 195 (53.7) | 6831 (25.3) | 4367 (48.2) |
| Four (V6) | 472 (58.0) | 1 (0.3) | 15028 (55.6) | 31 (0.3) |
| Five (V7) | 112 (13.8) | 0 (0.0) | 3051 (11.3) | 0 (0.0) |

| | | | | |
|---|---|---|---|---|
| **Interval in months between last dose and specimen collection,** median (interquartile range) | 2.5 (2-4) | 15.4 (14-17) | 3.3 (2-5) | 16.3 (15-18) |
| **2023/24 seasonal influenza vaccination** (≥14 days before specimen collection) | 651 (80.0) | 65 (17.9) | 21431 (79.3) | 2164 (23.9) |

Abbreviations: V3 to V7, total number of vaccine doses 3 to 7; MV/BV, monovalent or bivalent booster dose received between July and December 2022; IQR, interquartile range

[a] At least two chronic conditions among the following: chronic respiratory disease, hypertension, cardiovascular disease, neurological disorder, anaemia, diabetes, hypothyroidism, fluid and electrolyte disorders, cancer, kidney disease, obesity, psychosis, liver disease, coagulopathy, weight loss, drug abuse, alcohol abuse, ulcer, paralysis.

Note: Numbers are n and column proportions except otherwise indicated

**Supplementary Table 3**. Characteristics of study participants during KP period by outcome and vaccination status

| KP period | Cases | | Controls | |
|---|---|---|---|---|
| **Characteristic** | **XBB vaccinated** | **MV/BV vaccinated** | **XBB vaccinated** | **MV/BV vaccinated** |
| **N** | 781 | 204 | 15401 | 4299 |
| **Sex** | | | | |
| Female | 388 (49.7) | 106 (52.0) | 7949 (51.6) | 2142 (49.8) |
| Male | 393 (50.3) | 98 (48.0) | 7452 (48.4) | 2157 (50.2) |
| **Age, years** | | | | |
| Mean (standard deviation) | 81 (8.8) | 80 (8.7) | 79 (9.2) | 76 (9.3) |
| 60-69 | 90 (11.5) | 26 (12.7) | 2776 (18.0) | 1239 (28.8) |
| 70-79 | 221 (28.3) | 70 (34.3) | 5361 (34.8) | 1530 (35.6) |
| ≥80 | 470 (60.2) | 108 (52.9) | 7264 (47.2) | 1530 (35.6) |
| **Place of residence** | | | | |
| Home | 581 (74.4) | 174 (85.3) | 11577 (75.2) | 3680 (85.6) |
| Private homes for older people | 188 (24.1) | 26 (12.7) | 3466 (22.5) | 520 (12.1) |
| Other | 12 (1.5) | 4 (2.0) | 358 (2.3) | 99 (2.3) |
| **Chronic conditions** | | | | |
| At least two chronic conditions [a] | 621 (79.5) | 148 (72.5) | 11502 (74.7) | 2925 (68.0) |
| Chronic heart disease | 399 (51.1) | 95 (46.6) | 7230 (46.9) | 1775 (41.3) |
| Chronic lung disease | 293 (37.5) | 61 (29.9) | 5890 (38.2) | 1515 (35.2) |
| Cancer | 169 (21.6) | 38 (18.6) | 3457 (22.4) | 866 (20.1) |
| Neurologic disease / dementia | 130 (16.6) | 24 (11.8) | 2128 (13.8) | 449 (10.4) |
| Obesity | 99 (12.7) | 21 (10.3 | 1698 (11.0) | 489 (11.4) |
| **Documented prior infection history** | 158 (20.2) | 28 (13.7) | 3614 (23.5) | 1034 (24.1) |
| **Interval in months between prior infection and specimen collection,** median (interquartile range) | 23.6 (19-29) | 25.3 (10-28) | 21.2 (15-28) | 15.8 (8-28) |
| **Number of booster doses received** | | | | |
| One (V3) | 2 (0.3) | 9 (4.4) | 183 (1.2) | 166 (3.9) |
| Two (V4) | 54 (6.9) | 91 (44.6) | 987 (6.4) | 2209 (51.4) |
| Three (V5) | 174 (22.3) | 104 (51.0) | 3620 (23.5) | 1913 (44.5) |
| Four (V6) | 406 (52.0) | 0 (0.0) | 8152 (52.9) | 11 (0.3) |
| Five (V7) | 145 (18.6) | 0 (0.0) | 2459 (16.0) | 0 (0.0) |

| | | | | |
|---|---|---|---|---|
| **Interval in months between last dose and specimen collection,** median (interquartile range) | 8.0 (7-9) | 20.9 (20-22) | 7.2 (6-8) | 20.6 (20-22) |
| **2023/24 seasonal influenza vaccination** (≥14 days before specimen collection) | 634 (81.2) | 52 (25.5) | 12004 (77.9) | 1016 (23.6) |

Abbreviations: V3 to V7, total number of vaccine doses 3 to 7; MV/BV, monovalent or bivalent booster dose received between July and December 2022; IQR, interquartile range

[a] At least two chronic conditions among the following: chronic respiratory disease, hypertension, cardiovascular disease, neurological disorder, anaemia, diabetes, hypothyroidism, fluid and electrolyte disorders, cancer, kidney disease, obesity, psychosis, liver disease, coagulopathy, weight loss, drug abuse, alcohol abuse, ulcer, paralysis.

Note: Numbers are n and column proportions except otherwise indicated

**Supplementary Table 4.** XBB-vaccine effectiveness against COVID-19 hospitalization by time since XBB vaccination and subvariant period

| | **XBB period** | **JN period** | **KP period** |
|---|---|---|---|
| | Adjusted VE (%) (95% CI) | Adjusted VE (%) (95% CI) | Adjusted VE (%) (95% CI) |
| Global | 54.4 (45.9 to 61.6) | 22.7 (12.1 to 32.0) | 0.5 (-16.7 to 15.2) |
| **Time since last XBB-vaccination** | | | |
| 7-30d (month 1) | 53.2 (43.5 to 61.3) | 27.8 (3.4 to 46.0) | 67.4 (34.7 to 83.7) |
| 31-60d (month 2) | 60.4 (46.0 to 70.9) | 22.5 (6.2 to 36.0) | 57.2 (29.3 to 74.1) |
| 61-91d (month 3) | NE | 27.6 (14.5 to 38.8) | 31.0 (-6.0 to 55.0) |
| 92-121d (month 4) | NE | 19.8 (0.2 to 35.5) | 35.7 (-12.9 to 63.4) |
| 122-152d (month 5) | NE | 23.1 (-3.1 to 42.6) | 44.7 (-19.6 to 74.5) |
| 153-182d (month 6) | NE | 6.5 (-44.1 to 39.3) | 3.4 (-40.4 to 33.5) |
| 183-213d (month 7) | NE | -5.2 (-159.5 to 57.4) | 11.9 (-16.3 to 33.2) |
| 214-243d (month 8) | NE | NE | -2.1 (-26.8 to 17.9) |
| 244-274d (month 9) | NE | NE | -19.4 (-47.3 to 3.2) |
| 275-304d (month 10) | NE | NE | -14.2 (-49.2 to 12.7) |

Abbreviations: CI, confidence interval; d, days; NE, not estimable; VE, vaccine effectiveness.

Note: Logistic regression model comparing XBB-vaccinated with participants last vaccinated with ancestral monovalent or bivalent vaccines from July to December 2022 and adjusted for sex, age group, chronic conditions, place of residence and epi-week (2-week periods)

**Supplementary Table 5.** XBB-vaccine effectiveness against COVID-19 hospitalization restricted to participants with prior NAAT-confirmed infection, by subvariant period

| Subvariant period and comparison groups | Cases | Controls | Unadjusted VE (95% CI) | Adjusted VE (95% CI) |
|---|---|---|---|---|
| **Global (W2023-44 to W2024-33)** | | | | |
| XBB-vaccinated and prior infection | 378 | 11033 | | |
| vs MV/BV-vaccinated in 2022 regardless prior infection status | 1356 | 20346 | 48.6 (42.3 to 54.2) | 43.4 (35.9 to 50.1) |
| vs MV/BV-vaccinated in 2022 with documented prior infection | 216 | 3906 | 38.0 (26.5 to 47.8) | 28.2 (13.6 to 40.3) |
| **XBB period** | | | | |
| XBB-vaccinated and prior infection | 29 | 752 | | |
| vs MV/BV-vaccinated in 2022 regardless prior infection status | 556 | 4386 | 69.6 (55.5 to 79.2) | 73.6 (61.1 to 82.1) |
| vs MV/BV-vaccinated in 2022 with documented prior infection | 103 | 931 | 65.1 (46.8 to 77.2) | 65.5 (46.8 to 77.6) |
| **JN period** | | | | |
| XBB-vaccinated and prior infection | 146 | 5811 | | |
| vs MV/BV-vaccinated in 2022 regardless prior infection status | 363 | 9070 | 37.2 (23.7 to 48.3) | 41.1 (27.5 to 52.1) |
| vs MV/BV-vaccinated in 2022 with documented prior infection | 53 | 1747 | 17.2 (-13.9 to 39.8) | 14.1 (-18.8 to 37.9) |
| **KP period** | | | | |
| XBB-vaccinated and prior infection | 145 | 3104 | | |
| vs MV/BV-vaccinated in 2022 regardless prior infection status | 204 | 4299 | 1.6 (-22.4 to 20.8) | 5.9 (-19.3 to 25.8) |
| vs MV/BV-vaccinated in 2022 with documented prior infection | 23 | 693 | -10.8 (-120.2 to 10.0) | -24.9 (-97.3 to 21.0) |

Abbreviations: CI, confidence interval; MV/BV, ancestral monovalent or bivalent booster dose received between July and December 2022; NAAT, nucleic acid amplification test; VE, vaccine effectiveness; W, week

Note: Logistic regression model comparing XBB-vaccinated with participants last vaccinated with ancestral monovalent or bivalent vaccines from July to December 2022 and adjusted for sex, age group, chronic conditions, place of residence and epi-week

**Supplementary Table 6.** XBB-vaccine effectiveness against COVID-19 hospitalization, by age group and subvariant period

| | Age group | | |
|---|---|---|---|
| | **60-69-year-olds** | **70-79-year-olds** | **≥80-year-olds** |
| | Adjusted VE (%) (95% CI) | Adjusted VE (%) (95% CI) | Adjusted VE (%) (95% CI) |
| **Global** | 3.9 (-17.7 to 21.5) | 33.4 (24.1 to 41.5) | 33.5 (26.2 to 40.0) |
| **XBB period** | | | |
| Any time since XBB-vaccination | 47.5 (10.5 to 69.2) | 56.1 (40.6 to 67.6) | 54.9 (43.3 to 64.1) |
| 7-60d since XBB-vaccination | 47.5 (10.5 to 69.2) | 56.1 (40.6 to 67.6) | 54.9 (43.3 to 64.1) |
| **JN period** | | | |
| Any time since XBB-vaccination | -6.7 (-47.5 to 22.8) | 25.5 (7.5 to 40.1) | 28.8 (14.5 to 40.7) |
| 7-60d since XBB-vaccination | -23.7 (-91.7 to 20.2) | 34.7 (11.1 to 52.0) | 27.2 (6.2 to 43.6) |
| **KP period** | | | |
| Any time since XBB-vaccination | -47.2 (-130.3 to 5.9) | 10.3 (-18.5 to 32.0) | 6.6 (-16.3 to 25.0) |
| 7-60d since XBB-vaccination | 53.2 (-136.2 to 90.7) | 55.9 (-7.8 to 82.0) | 63.0 (28.3 to 77.9) |

Abbreviations: CI, confidence interval; d, days; VE, vaccine effectiveness.

Note: Logistic regression model comparing XBB-vaccinated participants by number of XBB doses (global or at two-month intervals from vaccination) with participants last vaccinated with ancestral monovalent or bivalent vaccines from July to December 2022, and adjusted for sex, chronic conditions, place of residence and epi-week (2-week periods)
